# Increasing access to certified nurse-midwives prevents use of medical interventions during labor: an application of g-computation and target trial emulation

**DOI:** 10.1101/2024.11.05.24316755

**Authors:** Elizabeth Simmons, Anna Austin, Mollie Wood, Alyssa J. Mansfield, Karen Sheffield-Abdullah, Kavita Singh

**Author notes:** Corresponding Author: Elizabeth Simmons.

## Abstract

**Background:** Prenatal care (PNC) led by a certified nurse-midwife (CNM) may reduce medical interventions and complications during labor and delivery, compared to physician-led care. We estimated the prevalence of six outcomes if we intervened to increase the number of low-risk pregnant people enrolled in PNC with a CNM.

**Methods:** We used 2014-2019 Pregnancy to Early Life Longitudinal data. The study population comprised people aged 18-55 with a low-risk pregnancy who initiated PNC with a physician or CNM at <13 weeks gestation and had a live birth. We used g-computation to estimate the prevalence and prevalence difference of cesarean sections, labor inductions, epidural use, postpartum hemorrhage, maternal infection and obstetric trauma under hypothetical scenarios where 10%, 20% and 50% more people and 100% more people with government-funded insurance had CNM-led PNC. We adjusted for payer of delivery, maternal age, education, race, and estimated 95% confidence intervals (CI) using bootstrap resampling.

**Results:** Among 130,835 pregnant people, one-fourth had CNM-led PNC. A 10% increase in CNM-led PNC resulted in a reduction in the prevalence of cesarean sections of -0.39 percentage-points (95% CI: 0.41 to -0.35), in labor inductions of -0.21 percentage-points (95% CI: -0.25 to -0.17), in epidural use of -0.85 percentage-points (95% CI: -0.89 to -0.80), and in maternal infections of-0.08 percentage-points (95% CI: -0.11 to -0.06), but an increase in prevalence of postpartum hemorrhage of 0.03 percentage-points (95% CI: 0.01 to 0.05) and no change in prevalence of obstetric trauma (0.00 percentage-points; 95% CI: -0.02 to 0.03). The effect estimates were larger for 20% and 50% increases.

**Conclusions:** A scale-up of CNM-led care contributes to a decrease in use of some medical interventions and complications during labor and delivery.

**Key Points:** Our results suggest that interventions to increase the number of low-risk pregnancies attended by CNMs versus physicians could reduce the use of some medical interventions and complications during labor and delivery.

With 10% more low-risk pregnant individuals enrolled in prenatal care with a CNM, the prevalence of cesarean sections, labor inductions, epidural analgesia, and maternal infection decreased, while the prevalence of postpartum hemorrhage increased slightly, and the prevalence of obstetric trauma did not change.

As barriers to accessing care with a CNM still exist for low-risk pregnant populations, policy makers and clinical providers should help to reduce barriers to access to CNMs to help improve clinical outcomes.

## Introduction

The rate of cesarean sections in the United States is alarmingly high and has been for decades. From 1996 to 2009, the cesarean section rate increased by 60%, from 20% to 33%, and has remained above 30% ever since.[1] Although an important medical procedure under certain circumstances, when used in a non-medically indicated way, cesarean sections can increase risks of harm, while providing limited medical benefit.[2] People who have a cesarean delivery are at increased risk of morbidities, such as uterine rupture, infection, and hemorrhage and mortality.[3] Healthy People 2030 has set a target of reducing the percentage of low-risk women who have not had a previous cesarean section to 23.6% (current estimates suggest this number is 26.3%).[4] There is an urgent need to address issues in the healthcare system that contribute to overuse of medical interventions, such as cesarean sections, and subsequent poor perinatal health outcomes.

A major barrier to improving perinatal health outcomes in the United States is a lack of access to midwives.[5] The majority of midwives in the United States are certified nurse-midwives (CNMs),[6] who are registered nurses that have additional training in midwifery to oversee care from menarche to menopause, including of pregnant populations. CNMs typically provide labor and delivery care to low-risk or healthy pregnant populations,[7] although they also co-manage care for higher risk pregnancies along with physicians. About 13% of pregnant people see a midwife for prenatal care (PNC) or as a delivery attendant (8% as a prenatal provider only and 11% as the primary birth attendant).[8] Most women who initiate PNC with a midwife continue midwifery care throughout the prenatal period and labor and delivery, with 5% transferring to a physician during pregnancy and an additional 20% transferring to a physician during labor and delivery.[9] Transfers during the prenatal period usually occur due to developing a high-risk complication that necessitates specialized care, while almost two-thirds of transfers during labor occur due to the need for a cesarean section.[9] Importantly, low-risk pregnant people who undergo prenatal care with CNMs are less likely to experience a medical intervention or complication during labor and delivery.[10–19]

In this study, we sought to quantify the number of medical interventions and labor/delivery complications that could be prevented under hypothetical scenarios where the use of CNMs as prenatal care providers increased for pregnant individuals in Massachusetts. Our study had two aims: (1) to assess the change in prevalence of cesarean sections, labor inductions, epidural analgesia, postpartum hemorrhage, obstetric trauma and maternal infection if 10%, 20% and 50% more low-risk pregnant people in Massachusetts were to enroll in prenatal care with a CNM; and (2) to assess the change in prevalence of the six outcomes if all low-risk pregnancies with government funded prenatal care in Massachusetts were to enroll in prenatal care with a CNM. As Medicaid covers 40% of all deliveries in the United States,[20] increasing access to midwifery-care for this population is particularly important. In addition, Medicaid enrollees are typically less healthy and are at higher risk of maternal morbidity or death compared with those who are commercially insured.[21,22]

## Methods

### Data and Sample

We used data from the Pregnancy to Early Life Longitudinal (PELL) Data System in Massachusetts for this study. PELL is a data source that links hospital discharge records and vital statistics data starting in 1999. Records are linked using a probabilistic matching program that uses variables such as facility code, medical record number, date of birth, sex, zip code, and birth weight.[23]

We used a target trial emulation framework to determine study eligibility and define key variables. This framework allowed us to use observational data to emulate an ideal randomized control trial (RCT) by outlining seven key components of a target trial protocol to emulate in an observational study design: eligibility, treatment strategies being compared, the assignment procedure, the follow-up period, outcomes of interest, causal contrasts of interest, and the analysis plan for the target trial (Table 1).[24]

**Table 1.** Applying Hernan’s Trial Emulation framework.

| Criteria | Target Trial | Observational Emulation |
| --- | --- | --- |
| **Eligibility** | • Currently pregnant,<br>• Initiates prenatal care by 12 6/7 weeks gestation with a viable pregnancy,<br>• Age 18-55 at last menstrual period,<br>• Low risk pregnancy, defined as not having any preexisting conditions or complications of previous pregnancies that would preclude initiating prenatal care with a midwife. | • Live birth,<br>• Initiates prenatal care by 12 6/7 weeks gestation,<br>• Age 18-34 at delivery,<br>• Initiate prenatal care in 2014-2019 in Commonwealth of Massachusetts,<br>• Low-risk pregnancy, defined as those without:<br>○ preconception conditions (diabetes, hypertension, cardiac disease, acute or chronic lung disease, renal disease, advanced maternal age (>35 years) and obesity (BMI>30))<br>○ or complications of previous pregnancies (preterm birth, small for gestational age infant, stillbirths, perinatal death or cesarean section) |
| **Treatment strategies being compared** | Treatment: Initiate prenatal care with a midwife<br>Control: Initiate prenatal care with a physician | Treatment: List midwives as at least one type of provider of prenatal care<br>Control: List OB-GYN (and not midwives) as provider type of prenatal care |
| **Assignment Procedure** | Participants are randomly assigned when entering prenatal care.<br>Participants will not be blinded to treatment arm assignment. | Participants choose treatment strategy.<br>Participants will not be blinded to treatment arm assignment. |
| **Follow-up period** | Randomization until 1-year post-partum, death or loss to follow-up (whichever comes first) | First prenatal visit until 6 weeks post-partum. |
| **Outcomes of interest** | use of cesarean section, labor induction, or epidural analgesia at delivery<br>occurrence of postpartum hemorrhage (within 24 hours and 12 | use of cesarean section, labor induction, or epidural analgesia at delivery<br>occurrence of postpartum hemorrhage (within 24 hours and 6 |
|  | weeks of delivery), obstetric trauma (within 12 weeks of delivery), or maternal infection (within 12 weeks of delivery) | weeks of delivery), obstetric trauma (within 6 weeks of delivery), or maternal infection (within 6 weeks of delivery) |
| **Causal contrasts of interest** | Prevalence difference | Prevalence difference |
| **Analysis plan** | Intention-to-treat: dichotomous exposure; treatment group to which participants are assigned. | Intention-to-treat: dichotomous exposure: treatment strategy of participant's choosing. |

Our study included all live births in Massachusetts between 2015 and 2019 to people ages 18-34 years with a low-risk pregnancy, who initiated prenatal care in the first trimester with a CNM or physician. We defined low-risk pregnancies as those without preconception conditions (diabetes, hypertension, cardiac disease, acute or chronic lung disease, renal disease, advanced maternal age (>35 years) and obesity (BMI>30)) or complications of previous pregnancies (preterm birth, small for gestational age infant, stillbirths, perinatal death or cesarean section) listed on the birth certificate. We made the decision to exclude these conditions/complications due to the fact that typically these conditions will make a person ineligible for CNM led prenatal care. [19,25–31] For study aim 2, we restricted to live births to people who listed a government payer source for prenatal care (i.e. Medicare, Medicaid/MassHealth, etc.). The institutional review board at UNC Chapel Hill reviewed this study and considered it exempt.

### Study Variables

The exposure for this study was prenatal care provider type (CNM vs. physician, which included obstetricians, family practitioners and other types of Medical Doctors (MD)). We defined the exposure based on a question on the Massachusetts Certificate of Live Birth that asks for the birth registrar to list all practitioner types an individual saw throughout prenatal care. Using the target trial emulation framework, our observation study emulated a target trial where individuals with low-risk pregnancies are randomized to either a CNM or physician provider when initiating prenatal care in the first trimester. Our analysis plan did not allow for individuals to change exposure groups, akin to an intention-to-treat analysis in an RCT setting. We did this to ensure that the baseline risk was similar among the exposure groups. As patients transfer from a lower to higher level of obstetric care (i.e. from a CNM to an obstetrician or maternal-fetal medicine specialist),[9] we identified those who initiated prenatal care with a CNM as any birth record that listed CNM as a practitioner type. We identified those who initiated prenatal care with a physician as those that listed an MD, but not a CNM, as a prenatal care practitioner type.

We examined six outcomes (Supplementary Table 1): cesarean delivery, labor induction, epidural, postpartum hemorrhage, obstetric trauma, and maternal infection. We identified cesarean deliveries, labor inductions and use of epidurals through birth certificate data. Using International Classification of Diseases, Ninth (ICD-9) and Tenth Revision (ICD-10) diagnostic codes, we identified individuals who had a postpartum hemorrhage (third-stage hemorrhage, other postpartum hemorrhage including atony, delayed/secondary postpartum hemorrhage), an obstetric trauma [major perineal laceration (third- and fourth-degree perineal lacerations, vulvar and perineal hematoma); other obstetric trauma (includes inversion of uterus, cervical laceration, high vaginal laceration, other injury to pelvic organs, joints, or ligaments, pelvic hematoma); ruptured uterus, and/or a maternal infection [genitourinary infection (pyelonephritis, urinary tract infection); amnionitis; other infection (unspecified pneumonia, unspecified bacterial infection, abscess); fever (maternal pyrexia during labor, unspecified); major puerperal infection (includes endometritis, sepsis, cellulitis, peritonitis, salpingitis); pyrexia of unknown origin in the puerperium; sepsis (generalized infection/septicemia during labor)]. We identified these outcomes in the 6 weeks following delivery, as some Medicaid enrollees become ineligible for Medicaid at this point. For postpartum hemorrhage, we also identified outcomes that occurred within 24 hours of delivery to capture primary postpartum hemorrhage.[32] We used a forward-backward mapping method to create the list code lists to identify these outcomes, using previously published ICD-9 diagnostic codes (Supplementary Table 2).[33–35] We also included three secondary outcomes to assess the impact of increases in CNM-led PNC on neonatal outcomes. We assessed the prevalence of preterm birth (<37 weeks gestation), having a low birth weight (<2500 grams) infant and having an Apgar score of <7 after 5 minutes, using birth certificate data.

For study aim 2, we identified people who used government-funded insurance as the prenatal care payer using the Massachusetts Certificate of Live Birth, including Medicare, Medicaid, MassHealth, CommonHealth, Commonwealth Care, Military health insurance (Champus, Tricare VA, etc.), and the Indian Health Service.

We identified potential confounders by creating a directed acyclic graph (DAG) (Supplementary Figure 1). Using the DAG, we identified the following minimally sufficient adjustment set: maternal age (18-24, 25-29, 30-34 years), maternal education (less than high school, high school/GED, some college, at least college graduate), maternal race (white, Black, Asian/Pacific Islander, Other) and payer of delivery (government-funded vs. other; study aim 1 only). Because maternal outcomes and choice of prenatal care provider can vary depending on number of previous pregnancies, we included parity (nulliparous vs. primi-/multiparous) as an effect measure modifier using stratified models in a sensitivity analysis.

### Statistical Analysis

We used the non-parametric g-formula to assess the change in prevalence of the six outcomes under four hypothetical scenarios: a 10%, 20% and 50% increase in use of CNMs as prenatal care providers among the population of all low-risk live births, and use of CNMs as prenatal care providers for all low-risk live births among government-funded pregnancies. The g-formula has been described in detail elsewhere.[36,37] Briefly, like other causal inference methods, the g-formula relies on estimation of unobserved counterfactual outcomes that correspond to hypothetical exposures, in addition to the observed outcomes that correspond to true exposures experienced in a study. To implement this methodology, we used a two-step process.[38,39] First, we assessed the prevalence of each outcome using a linear binomial model, specified as follows: *P*(*Outcome*^*TE*^ = 1) = *β*_0_ + *β*_1_*CNM*^*TE*^ + *β*_2_*W*, where *Outcome^TE^* is one of the six outcomes under the true exposure, *CNM^TE^* is the true exposure and *W* is vector that includes all confounders. Then, we used this regression model to predict two counterfactual outcomes for each live birth, given *W:* the outcome had that individual enrolled in CNM-led PNC and the outcome had than individual enrolled in physician-led PNC. For the group that enrolled in prenatal care with a provider that was not a CNM (i.e., a physician), we changed this exposure to CNM with a probability of 0.1, 0.2, 0.5 and 1 for a 10%, 20%, 50% and 100% increase (among live births with government-funded insurance only) in use of CNMs, respectively. We estimated prevalence under each scenario and the prevalence difference of each scenario compared to the true exposure and estimated 95% confidence intervals (CI) using bootstrap resampling.[40]

## Results

Our final sample included 130,835 pregnancies. From the total sample of all live births (N= 339,377) between 2015 and 2019, we excluded 2,225 pregnancies (1%) to people <18 or >55 years, 176,407 pregnancies (52%) that were not low-risk, 28,147 pregnancies (8%) that initiated PNC after the first trimester, and 1,763 pregnancies (1%) that did not see a CNM or physician for prenatal care (Figure 1). For study objective 2, we additionally excluded 87,277 live births to pregnant people who did not have government-funded insurance, with a final sample of 43,558 live births.

**Figure 1.**
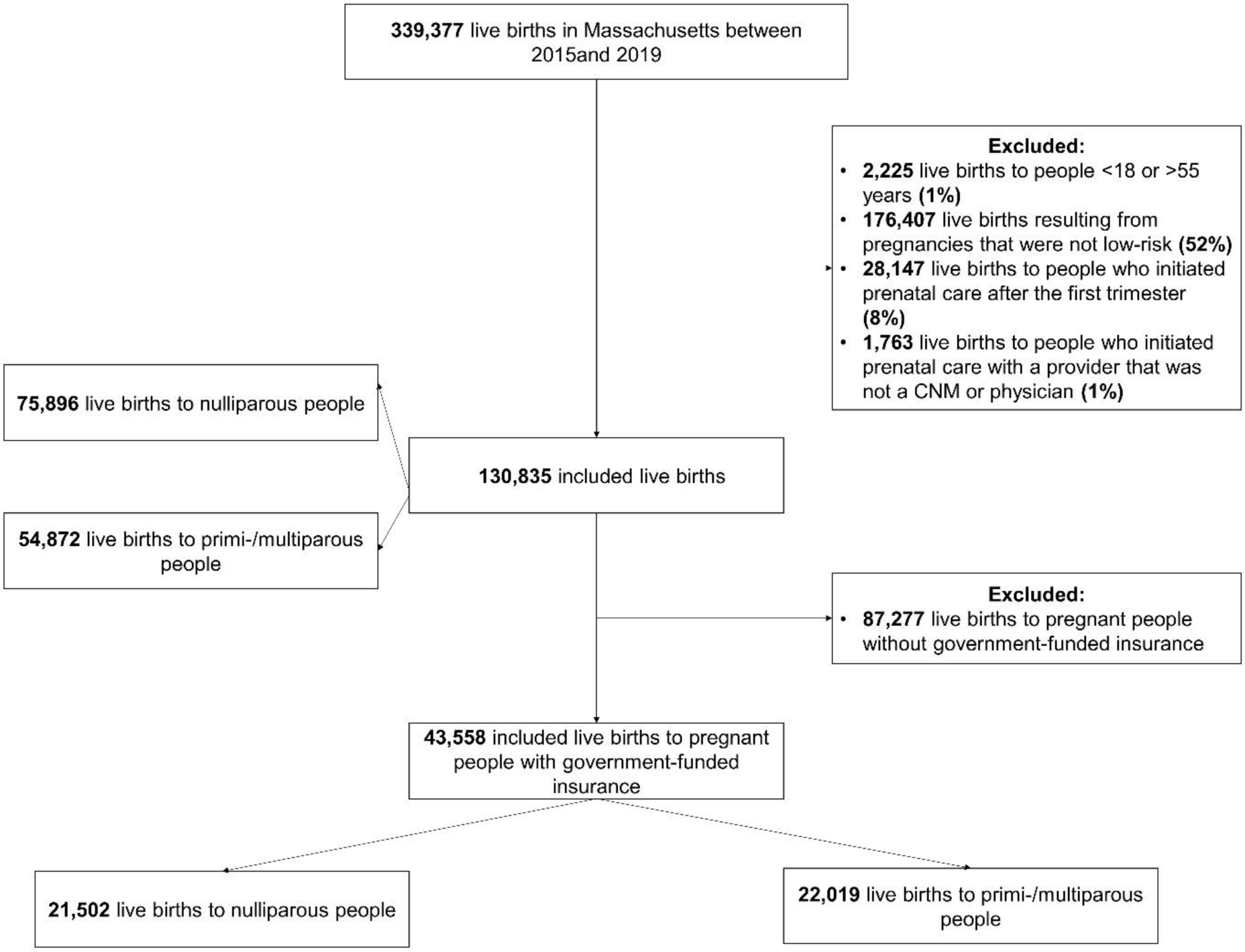
Study Flow Diagram.

One in four pregnant people initiated PNC with a CNM. Among those who initiated prenatal care with a CNM, 15.0% had a cesarean delivery, 20.2% had a labor induction, 66.5% had an epidural, 5.8% had a postpartum hemorrhage, 5.1% had an obstetric trauma and 5.8% had a maternal infection. Among those who initiated prenatal care with a physician, 20.6% had a cesarean delivery, 22.8% had a labor induction, 78.2% had an epidural, 5.4% had a postpartum hemorrhage, 5.2% had an obstetric trauma, and 7.1% had a maternal infection. People who initiated prenatal care with a CNM, on average, had fewer number of study outcomes than those who initiated with a physician. Those who initiated PNC with a CNM were slightly younger, had fewer years of education, were less likely to be nulliparous and more likely to have a government source as primary payer of prenatal care (Table 2). Pregnant people with CNM-led PNC had similar rates of pregnancy complications and delivery characteristics to those with physician-led PNC.

**Table 2.** Characteristics of full sample and by exposure group.

|  | Full Sample<br>N= 130,835<br>n (col%) | Missing<br>n | Prenatal Care<br>CNM<br>N= 30,824<br>n (col%) | Attendant<br>Physician<br>N= 100,011<br>n (col%) |
| --- | --- | --- | --- | --- |
| **Outcomes** |  |  |  |  |
| Cesarean section | 25,180 (19.3) | 3 | 4,608 (15.0) | 20,572 (20.6) |
| Labor induction | 28,973 (22.1) | 1 | 6,221 (20.2) | 22,752 (22.8) |
| Epidural analgesia | 98,682 (75.4) | 1 | 20,485 (66.5) | 78,197 (78.2) |
| Postpartum hemorrhage (within 24 hours) | 7,140 (5.5) | 827 | 1,749 (5.8) | 5,391 (5.4) |
| Postpartum hemorrhage (within 6 weeks) | 7,140 (5.5) | 827 | 1,749 (5.8) | 5,391 (5.4) |
| Obstetric trauma | 6,674 (5.1) | 827 | 1,538 (4.9) | 5,136 (5.2) |
| Maternal infection | 8,781 (6.8) | 827 | 1,752 (5.8) | 7,029 (7.1) |
| Number of outcomes |  |  |  |  |
| 0 | 21,894 (16.7) | 0 | 7,778 (25.2) | 14,116 (14.1) |
| 1 | 56,851 (43.5) |  | 12,597 (40.9) | 44,254 (44.3) |
| 2 | 39,802 (30.4) |  | 7,989 (25.9) | 31,813 (31.8) |
| ≥3 | 12,288 (9.4) |  | 2,460 (8.0) | 9,828 (9.8) |
| **Secondary Outcomes** |  |  |  |  |
| Preterm Birth | 9,514 (7.3) | 0 | 1,510 (4.9) | 8,004 (8.0) |
| Low birth weight | 8,423 (6.4) | 0 | 1,333 (4.3) | 7,090 (7.1) |
| Apgar score <7 at 5 minutes | 1,863 (1.4) | 214 | 371 (1.2) | 1,492 (1.5) |
| **Maternal Characteristics** |  |  |  |  |
| Age |  |  |  |  |
| 18-24 | 21,674 (16.6) | 0 | 6,079 (19.7) | 15,595 (15.6) |
| 25-29 | 40,941 (31.3) |  | 10,234 (33.2) | 30,707 (30.7) |
| 30-34 | 68,220 (52.1) |  | 14, 511 (47.1) | 53,709 (53.7) |
| Education |  |  |  |  |
| Less than high school | 8,796 (6.7) | 3,385 | 3,018 (10.0) | 5,778 (5.9) |
| High school / GED | 19,248 (14.7) |  | 5,230 (17.4) | 14,018 (14.4) |
| Some college | 30,484 (23.3) |  | 7,995 (26.5) | 22,489 (23.1) |
| At least a college graduate | 68,922 (52.7) |  | 13,893 (46.1) | 55,029 (56.6) |
| Race |  |  |  |  |
| White | 82,481 (63.0) | 2,366 | 19,137 (62.9) | 63,344 (64.6) |
| Black | 9,516 (7.3) |  | 2,268 (7.5) | 7,248 (7.4) |
| Asian/Pacific Islander | 14,863 (11.4) |  | 2,690 (8.8) | 12,173 (12.4) |
| Other | 21,609 (16.5) |  | 6,320 (20.8) | 15,289 (15.6) |
| Government-funded payer for prenatal care | 43,558 (33.3) | 1 | 12,853 (41.7) | 30,705 (30.7) |
| Nulliparous | 75,896 (58.0) | 67 | 16,557 (53.7) | 59,339 (59.4) |
| **Current pregnancy complications** |  |  |  |  |
| Gestational diabetes | 5,454 (4.2) | 0 | 1,344 (4.4) | 4,110 (4.1) |
| Gestational hypertension, pre-eclampsia, or eclampsia | 7,238 (5.5) | 0 | 1,533 (5.0) | 5,705 (5.7) |
| **Delivery characteristics** |  |  |  |  |
| Hospital delivery | 130,239 (99.5) | 2 | 30,455 (98.8) | 99,784 (99.8) |
| Non-vertex presentation | 4,326 (3.3) | 1 | 674 (2.2) | 3,652 (3.7) |
| Placenta previa | 599 (0.5) | 1 | 95 (0.3) | 504 (0.5) |
| Abruptio placenta | 1,013 (0.8) | 1 | 175 (0.6) | 838 (0.8) |
| Cord prolapse | 228 (0.2) | 1 | 25 (0.1) | 203 (0.2) |
| Prolonged labor (>20 hours) | 1,970 (1.5) | 1 | 433 (1.4) | 1,537 (1.5) |
| Electronic fetal monitoring | 119,351 (91.2) | 0 | 29,468 (95.6) | 89,883 (89.9) |

In Figure 2, we display the change in prevalence of the six primary outcomes as CNM-led PNC increases. Among the population of all live births following low-risk pregnancies, as CNM-led PNC increases 10%, 20% and 50% from the true exposure, the prevalence of cesarean sections (19.3% under true exposure vs. 17.3% with a 50% increase), labor induction (22.3% under true exposure vs. 21.2% with a 50% increase), epidural (75.4% under true exposure vs. 71.1% with a 50% increase), and maternal infection (6.7% under true exposure vs. 6.2% with a 50% increase) decreases. The prevalence of obstetric trauma remained the same under the different exposure regimens (5.2%) and the prevalence of postpartum hemorrhage increased slightly (5.2% under the true exposure vs. 5.6% with a 50% increase). Assigning all low-risk pregnancies enrolled in government-funded payment programs to initiate prenatal care with a CNM resulted in a decrease in prevalence for all six outcomes.

**Figure 2.**
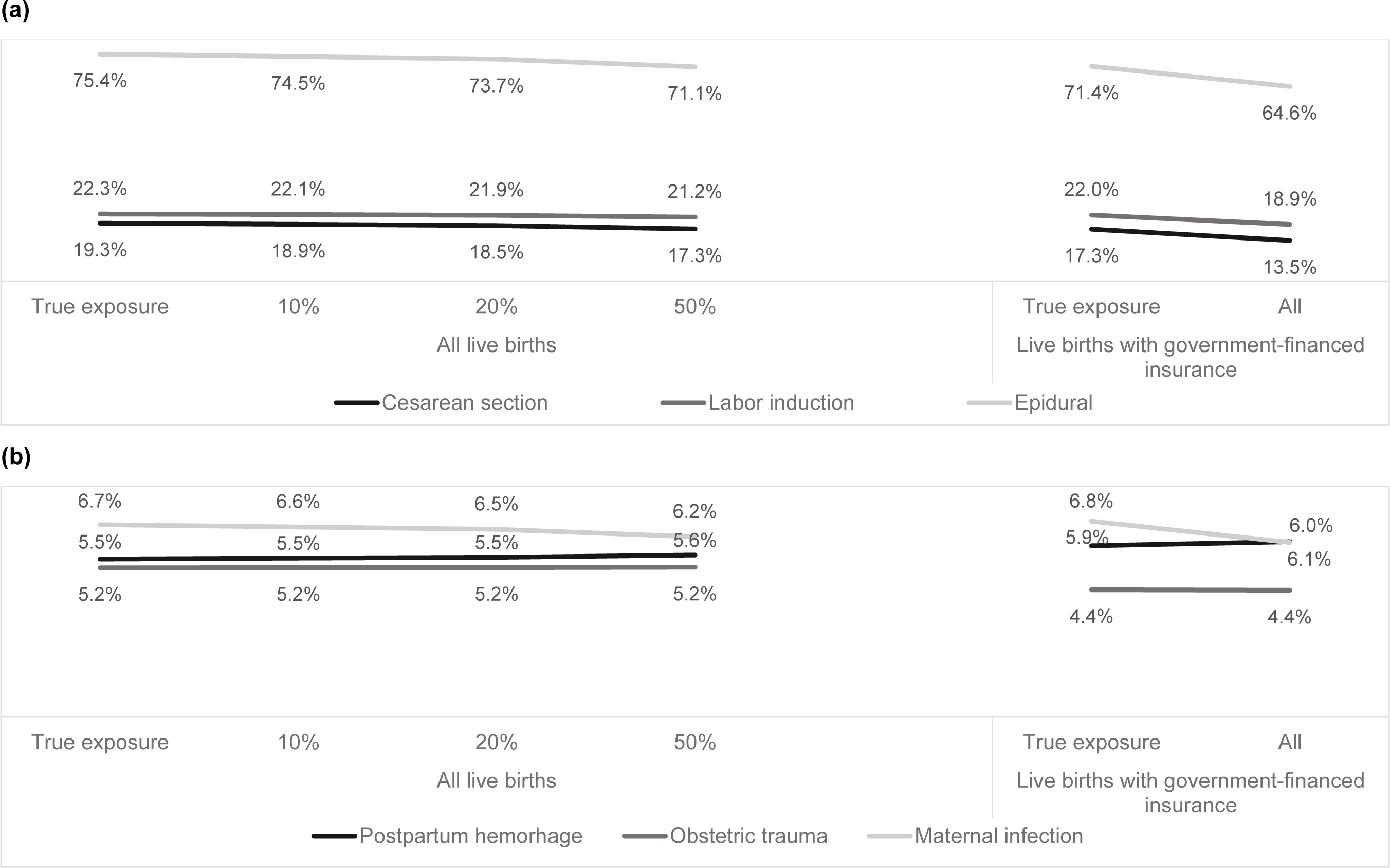
Change in prevalence of (a)cesarean section, labor induction, epidural and (b)postpartum hemorrhage, obstetric trauma, maternal infection with increase in use of CNMs as providers of prenatal care among low-risk pregnancies in Massachusetts, 2015-2019.

We show the prevalence difference in the six outcomes for an increase in CNM-led PNC compared to the true exposure in Figure 3. A 10% increase in CNM-led PNC resulted in a reduction in prevalence of cesarean sections of -0.39 percentage-points (95% CI: 0.41 to -0.35), in labor inductions of -0.21 percentage-points (95% CI: -0.25 to -0.17), in epidural use of -0.85 percentage-points (95% CI: -0.89 to -0.80), and in maternal infections of -0.08 percentage-points (95% CI: -0.11 to -0.06) among all live births to low-risk pregnancies. The prevalence difference of postpartum hemorrhage with a 10% increase in use of CNMs as prenatal care providers compared with the true exposure was 0.03 percentage-points (95% CI: 0.01 to 0.05).

**Figure 3.**
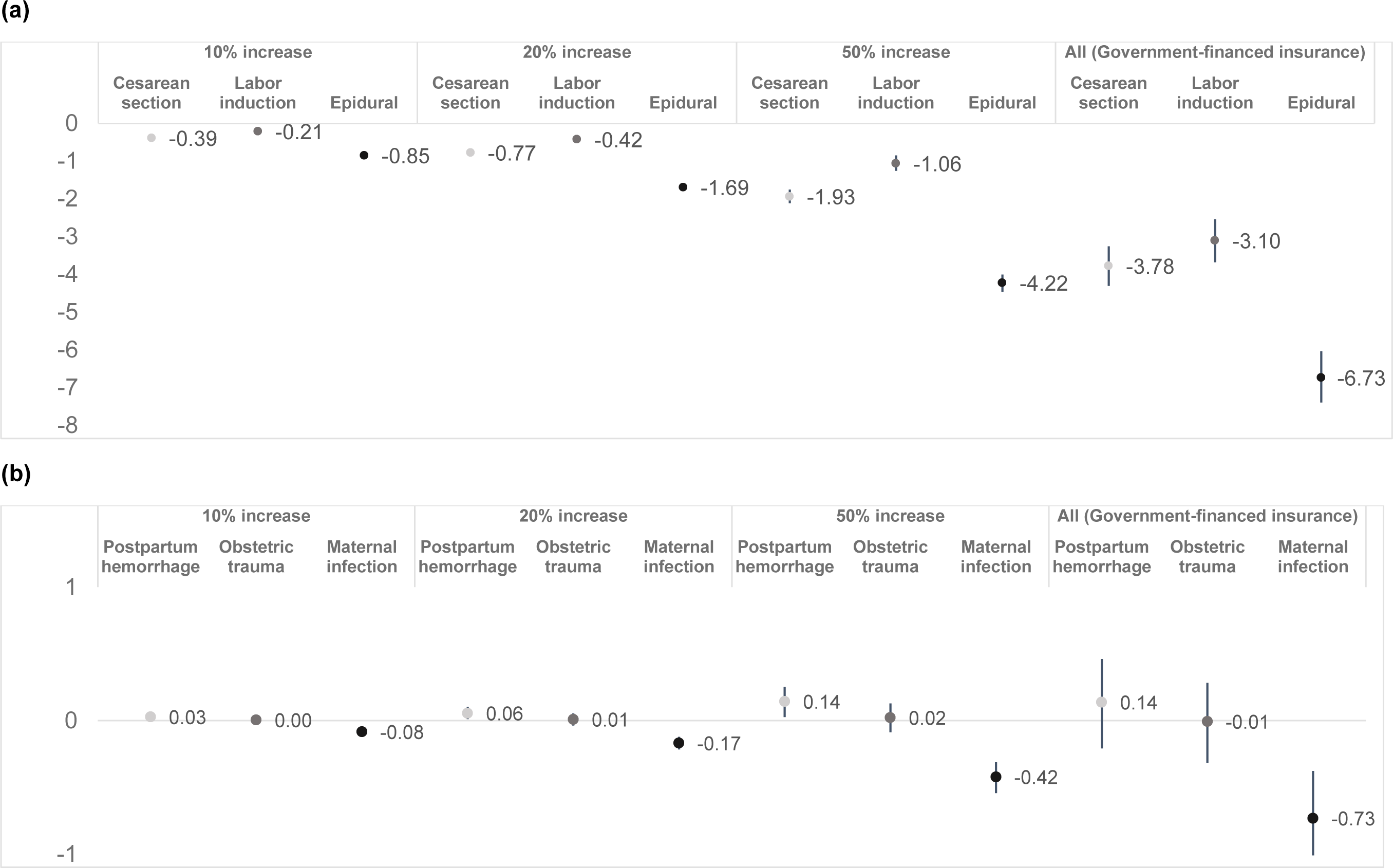
Prevalence difference of (a)cesarean section, labor induction, epidural and (b)postpartum hemorrhage, obstetric trauma, maternal infection comparing increase in use of CNMs as prenatal care providers with the true exposure among low-risk pregnancies in Massachusetts, 2015-2019.

A 20% and 50% increase in the use of CNMs as prenatal care providers resulted in stronger reductions in all outcomes, except postpartum hemorrhage, which increased. The use of CNMs for prenatal care providers for all low-risk pregnancies with government-funded insurance for prenatal care resulted in a decrease in prevalence of cesarean section, labor induction, epidural and maternal infection and no change in prevalence of postpartum hemorrhage and obstetric trauma. We observed a decrease in all secondary outcomes, including preterm birth, low infant birthweight, and Apgar score of <7 after 5 minutes, with increasing use of CNMs (Supplementary Figures 2 and 3).

When we stratified the analysis by parity (Supplementary Figures 4-6), we found that the prevalence difference of epidural use with increasing use of CNMs compared with the true exposure was greater among primi-/multiparous people than nulliparous people. In addition, we did not observe an increase in the prevalence of postpartum hemorrhage among primi-/multiparous people, but we did observe an increase in the prevalence of obstetric trauma with increasing use of CNMs. The prevalence differences of neonatal outcomes with increasing use of CNMs compared with the true exposure were similar among both groups.

## Discussion

Our study estimated the change in prevalence of medical interventions and complications during labor and delivery under hypothetical scenarios in which the use of CNMs as prenatal care providers increased among low-risk pregnancies. We found that the prevalence of cesarean sections, labor inductions, epidural analgesia and maternal infections decreased as the use of CNMs increased 10%, 20% and 50% among all low-risk pregnancies, while the prevalence of obstetric traumas stayed the same and the prevalence of postpartum hemorrhage increased slightly. When all low-risk pregnancies with government-funded insurance for prenatal care were assigned prenatal care with a CNM, we found that the prevalence of all six outcomes decreased.

The reduction in use of medical interventions during labor and delivery among pregnancies undergoing care with a CNM compared with a physician is well documented.[19,31,41,42] Because CNMs are educated regarding the natural physiologic process of labor and birth and trained to allow childbirth to proceed with minimal interventions, patients undergoing CNM-led prenatal and labor and delivery care are less likely to receive a cesarean section than those undergoing physician-led care.. CNMs also provide longer visits and increased prenatal support support,[43] which lead to healthier pregnancies that allow labor and delivery to progress with minimal intervention.

Labor inductions can occur for a multitude of reasons, including premature rupture of membranes, post-term pregnancy, or for elective (nonmedical) reasons.[2] The benefits and harms of elective labor inductions are a debated topic in the literature.[44] As CNMs are educated to allow labor to progress with minimal interventions,[45] CNM patients may be less likely to undergo labor inductions. Epidural analgesia is the most effective and requested form of pain management during labor and delivery,[46] but is also associated with increased risk of medical intervention during labor and delivery.[2,47] Many alternative pain management techniques exist, such as nitrous oxide, systemic opioids, water immersion and labor support from a trained doula,[2] but are often underutilized due to lack of education on behalf of both patients and providers.[48] However, some of these alternative forms of pain management might be more accessible in places where CNMs work.[48] A reduction in the use of epidural analgesia with increasing use of CNMs could reflect patient choice, as patients who want less intervention will likely choose a CNM as their prenatal care provider and labor and delivery attendant. It is unclear if the same reduction would be observed among a population that is assigned, rather than self-selected into, CNM-led care at initiation of prenatal care.

People who undergo a cesarean delivery are at higher risk of maternal infection,[49,50] while those undergoing instrumental vaginal deliveries are at a higher risk of major perineal lacerations (the main type of obstetric laceration).[51] As patients undergoing care with a CNM are less likely to undergo a cesarean section or an instrumental vaginal delivery, we hypothesized that prevalence of maternal infection and obstetric trauma would decrease as the use of CNMs increased. While we did observe a decrease in prevalence of maternal infections when use of CNMs was increased, we found that the prevalence of obstetric trauma did not change when CNM use was increased among the population of all live births. This scenario is a more extreme policy change than the 10%, 20% and 50% increase in use of CNMs we examined among live births, which could explain why we observed a change in this population and not among all live births.

Surprisingly, we found that the prevalence of postpartum hemorrhage increased slightly as the use of CNMs increased. To our knowledge, only one other study has observed an increase in prevalence of postpartum hemorrhage among patients who undergo prenatal care with a CNM.[19] While this may reflect a true increase, it may also reflect a difference in measurement and identification of postpartum hemorrhage across provider types. The increased risk of postpartum hemorrhage in CNM-led PNC care demonstrated by Weisband et al [19] and in our study may suggest that perhaps training on how to measure blood loss during labor and delivery differs between CNMs and physicians, or between vaginal and cesarean deliveries. In addition, guidance on estimating postpartum hemorrhage among vaginal deliveries changed in 2017, from loss of at least 500 mL to 1,000 mL of blood regardless of delivery type.[32] In a post hoc analysis, we assessed the impact of increasing CNM-led PNC prior to 2017 and in 2017 and later years. We found that the prevalence of postpartum hemorrhage did not increase with more CNM-led PNC prior to 2017, but did increase in 2017-2019 (Supplementary Figure 7). In addition, CNMs are less likely to perform active management of the third stage of labor,[52] which could increase prevalence of postpartum hemorrhage. Patients with CNM-led care are less likely to want interventions during labor and delivery, which could delay active management of postpartum hemorrhage. As a second post hoc analysis, we assessed the change in prevalence of severe postpartum hemorrhage (postpartum hemorrhage and blood transfusion) and found no change with increasing use of CNMs (Supplementary Figure 8). We think that these additional analyses suggest that our hypothesis regarding a CNM’s waiting to intervene in delivery helps to explain our findings. We did not observe an increase in prevalence of postpartum hemorrhage among those with previous pregnancies. We may be more easily able to identify a low-risk population among people with previous pregnancy history.

Importantly, we found that increasing the use of CNMs among this population resulted in reduced prevalence of preterm births, and infants of low birth weight and with an Apgar score of <7 at 5 minutes after delivery. CNMs typically have longer appointments with their prenatal patients and provide additional education and support during the prenatal period.[43] In addition, CNM patients often feel a sense of connection and trust with their providers, resulting in being more upfront about health conditions and prompt treatment of these conditions.[53] For these reasons, care with a CNM could result in healthier pregnancies and improved neonatal health outcomes.

Our study has some limitations. First, the sample in this study was restricted to live births, which could induce a type of collider bias (known as live birth bias) in our effect estimates.[54,55] Future work should replicate these analyses among pregnant populations that include outcomes other than live births. Second, there could be some misclassification of the attendant at prenatal care initiation. In our analysis, 7.6% of the sample (n=9,450) listed both a CNM and physician as a practitioner of prenatal care. We assumed that anyone with both a CNM and physician listed on the birth certificate as prenatal attendants were individuals who initiated care with a CNM and then transferred to care with a physician. However, these could have been individuals who saw both CNMs and physicians throughout their pregnancy. Third, there could be residual or uncontrolled confounding. We attempted to limit confounding through use of a DAG to identify a minimally sufficient adjustment set. Importantly, we could not control for a patient’s choice of prenatal care provider in our analysis. Fourth, we were unable to ascertain whether cesarean deliveries were medically indicated or emergent. Future efforts should help make this distinction as it is important to both make sure those individuals who need a cesarean are receiving one, but that those who do not have a medical indication are avoiding an unnecessary medical procedure. Fifth, there is some heterogeneity in the subpopulation of individuals who had government-financed insurance for prenatal care, as different types of people will have different types of insurance to finance their pregnancy care. Finally, these results may not be generalizable to pregnant populations outside of Massachusetts. Massachusetts has a unique healthcare environment, for example, adopting major healthcare reform prior to implementation of the Patient Care and Affordable Care Act (ACA). In addition, almost 20% of this population initiated prenatal care with a CNM, nearly twice that of other estimates.[8] Future work should replicate these analyses in different populations.

## Conclusion

CNMs are often underused providers trained to oversee care of low-risk pregnant populations. Our study demonstrates that increasing the use of CNMs as providers of prenatal care for low-risk pregnancies could help to prevent unnecessary medical interventions and resulting complications during labor and delivery, although caution is warranted given the small elevated risk of PPH. Major barriers to accessing CNMs still exist, including restrictions on how and where a CNM can practice and how CNMs are reimbursed for the care they provide. Policy makers and clinical providers could help to improve access to CNMs by removing requirements for CNMs to have supervising physicians or written collaborative agreements in order to practice, improving reimbursement policies for CNMs and ensuring that CNMs are eligible for hospital privileges.

## Data Availability

All data are available through the Massachusetts Department of Public Health.

