## Supplementary Materials for "Increasing access to certified nurse-midwives prevents use of medical interventions during labor: an application of g-computation and target trial emulation"

**Supplementary Table 1. Data source, definition and timing of variables.**

| Variable | Data Source | Definition | Timing |
|---|---|---|---|
| **Exposure** | | | |
| Provider type of prenatal care | Birth certificate | CNM vs. other | At prenatal care initiation |
| **Primary Outcomes** | | | |
| Cesarean section | Birth Certificate | Yes vs. no | At labor and delivery |
| Labor Induction | Birth Certificate | Yes vs. no | At labor and delivery |
| Epidural Analgesia | Birth Certificate | Yes vs. no | At labor and delivery |
| Postpartum hemorrhage | Hospital Discharge | Any postpartum hemorrhage vs. none | Within 24 hours and 6 weeks of delivery |
| Obstetric Trauma | Hospital Discharge | Any $3^{rd}/4^{th}$ degree laceration, $4^{th}$ degree laceration, other obstetric trauma, ruptured uterus vs. none | Within 6 weeks of delivery |
| Maternal Infection | Hospital Discharge | Any genitourinary infection, amnionitis, other infection, fever, major puerperal infection, postpartum fever of unknown origin, sepsis vs. none | Within 6 weeks of delivery |
| **Secondary Outcomes** | | | |
| Preterm Birth | Birth Certificate | Yes vs. no | At labor and delivery |
| Low birth weight | Birth Certificate | Yes vs. no | At labor and delivery |
| Apgar score $\geq$7 at 5 minutes | Birth Certificate | Yes vs. no | At labor and delivery |
| **Subgroup** | | | |

| Medicaid/MassHealth enrollees | Birth Certificate | Yes for prenatal care vs. no for prenatal care | During prenatal care |
|---|---|---|---|

**Supplementary Table 2. ICD-10 Codes for Outcomes**

| Outcome | Definition | ICD-9 Codes | ICD-9 Validity Metrics | ICD-9 Source | ICD-10 Codes | Source |
|---|---|---|---|---|---|---|
| Cesarean Section | | 74.0–74.2, 74.4, 74.99, 669.70*–669.71* | Sensitivity=100%; PPV=99.7% (98%-100%) | 152,172 | | |
| Labor Induction | • Induction of labor by artificial rupture of membranes | 73.01 | Sensitivity=63.5% (60.4%-66.5%); Specificity=93.0% (91.7%-94.1%); PPV=82.5% (79.5%-85.1%); NPV= 83.1% (81.4% - 84.6%) | 181 | | |
| | • Surgical induction of labor | 73.1 | | | | |
| | • Medical induction of labor | 73.4 | | | | |
| | • Other genitourinary instillation | 96.49 | | | | |
| Use of epidural analgesia | | 03.90 | | 182 | | |
| Postpartum Hemorrhage | • Postpartum hemorrhage (third-stage hemorrhage, other postpartum hemorrhage including atony, delayed/secondary postpartum hemorrhage) | All 666 | Sensitivity=92% (82%-97%) | 152,153 | All O72 | 154 |
| Obstetric trauma | • Major perineal laceration (third- and fourth-degree perineal lacerations, vulvar and perineal hematoma) | 664.2; 664.3; 664.5 | | 152 | O70.2; O70.3; O71.7 | |
| | • Other obstetric trauma (includes inversion of uterus, cervical laceration, high vaginal laceration, other | 665.2–665.9 | | | O71.2 - O71.7; O71.89, O71.9 | |

| Outcome | Definition | ICD-9 Codes | ICD-9 Validity Metrics | ICD-9 Source | ICD-10 Codes | Source |
|---|---|---|---|---|---|---|
| | injury to pelvic organs, joints, or ligaments, pelvic hematoma) | | | | | |
| | • Ruptured uterus | 665.0–665.1 | | | O71.9, O71.02, O71.03, O71.1 | |
| Maternal infection | • Genitourinary infection (pyelonephritis, urinary tract infection) | 646.6; 590; 599.0; 583.81 (added based on reverse coding ICD-10 codes N16 and N15.9) | | | O23.00, O23.10, O23.20, O23.30, O23.40, O23.519, O23.529, O23.599, O23.90, O23.91-O23.93, O86.11, O86.13, O86.19, O86.20 - O86.22; N11.0, N11.8, N10, N15.1, N28.84 - N28.86, N12, N16, N15.9; N39.0 | |
| | • Amnionitis | 658.4 | | | O41.1090, O41.1290, O41.1490, O41.1010, O41.1020, O41.1030, O41.1210, O41.1220, O41.1230, O41.1410, O41.1420, O41.1430 | |

| Outcome | Definition | ICD-9 Codes | ICD-9 Validity Metrics | ICD-9 Source | ICD-10 Codes | Source |
|---|---|---|---|---|---|---|
| | • Other infection (unspecified pneumonia, unspecified bacterial infection, abscess) | 486; all 041; 682 | | | J18.9; B95.0 – B95.8, B96.0 – B96.7, A49.3, B96.89, B96.81; K12.2, L03.211, L03.212, L03.213, L03.221, L03.222, L03.319, L03.329, L03.119, L03.129, L03.317, L03.811, L03.818, L03.891, L03.898, L03.90, L03.91 | |
| | • Fever (maternal pyrexia during labor, unspecified) | 659.2 | | | O75.2 | |
| | • Major puerperal infection (includes endometritis, sepsis, cellulitis, peritonitis, salpingitis) | 670; all 615 | | | O86.89, O86.12, O85, O86.81; all N71 | |
| | • Pyrexia of unknown origin in the puerperium | 672 | | | O86.4 | |
| | • Sepsis (generalized infection/septicemia during labor) | 659.3 | | | O75.3 | |

*excluded from calculation of validity metrics

**Supplementary Figure 1. Directed acyclic-graph**

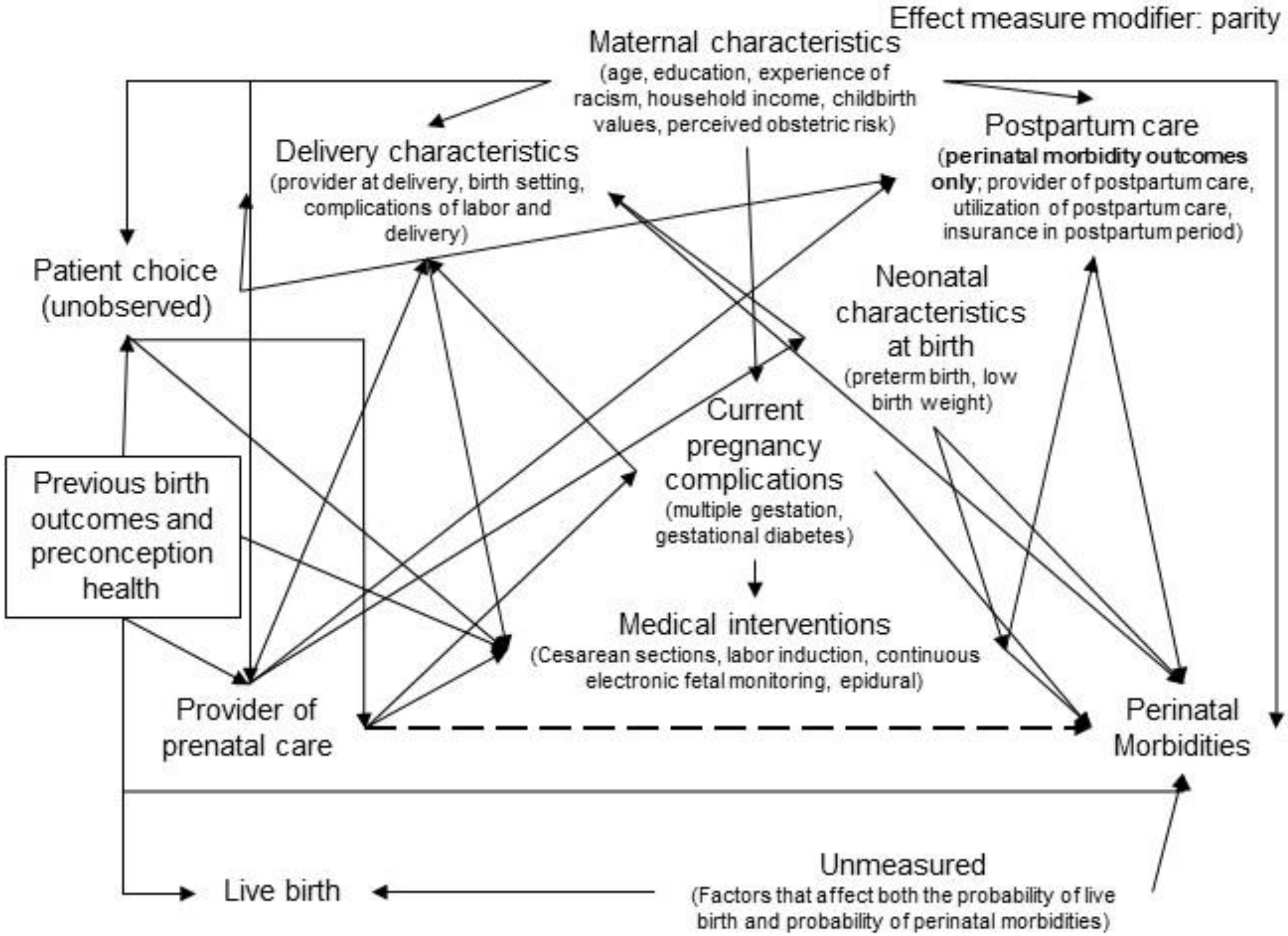

**Supplementary Figure 2. Change in prevalence of preterm birth, low birth weight infants and infants with Apgar score <7 with increase in use of CNMs as providers of prenatal care among low-risk pregnancies in Massachusetts, 2015-2019**

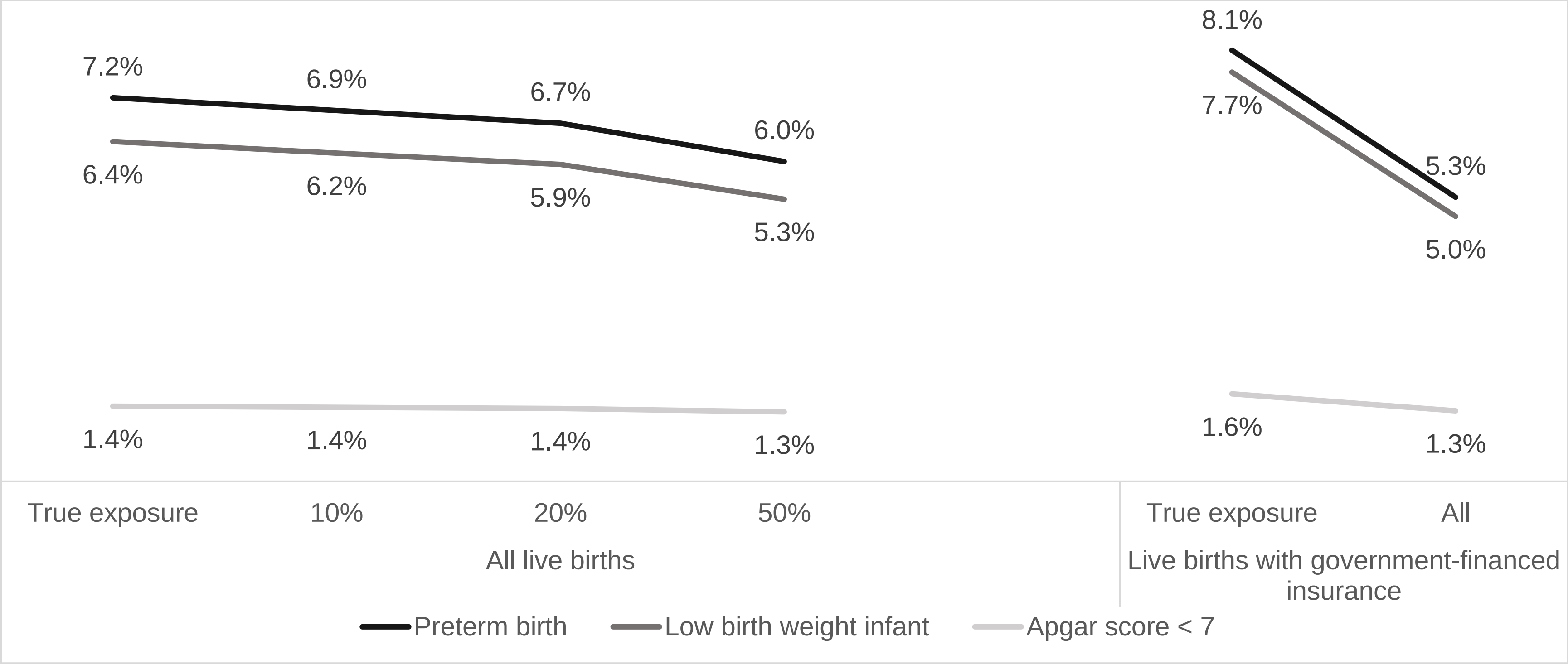

**Supplementary Figure 3. Prevalence difference of preterm birth, low birth weight infants and infants with Apgar < 7 comparing increase in use of CNMs as prenatal care providers with natural course among low-risk pregnancies in Massachusetts, 2015-2019**

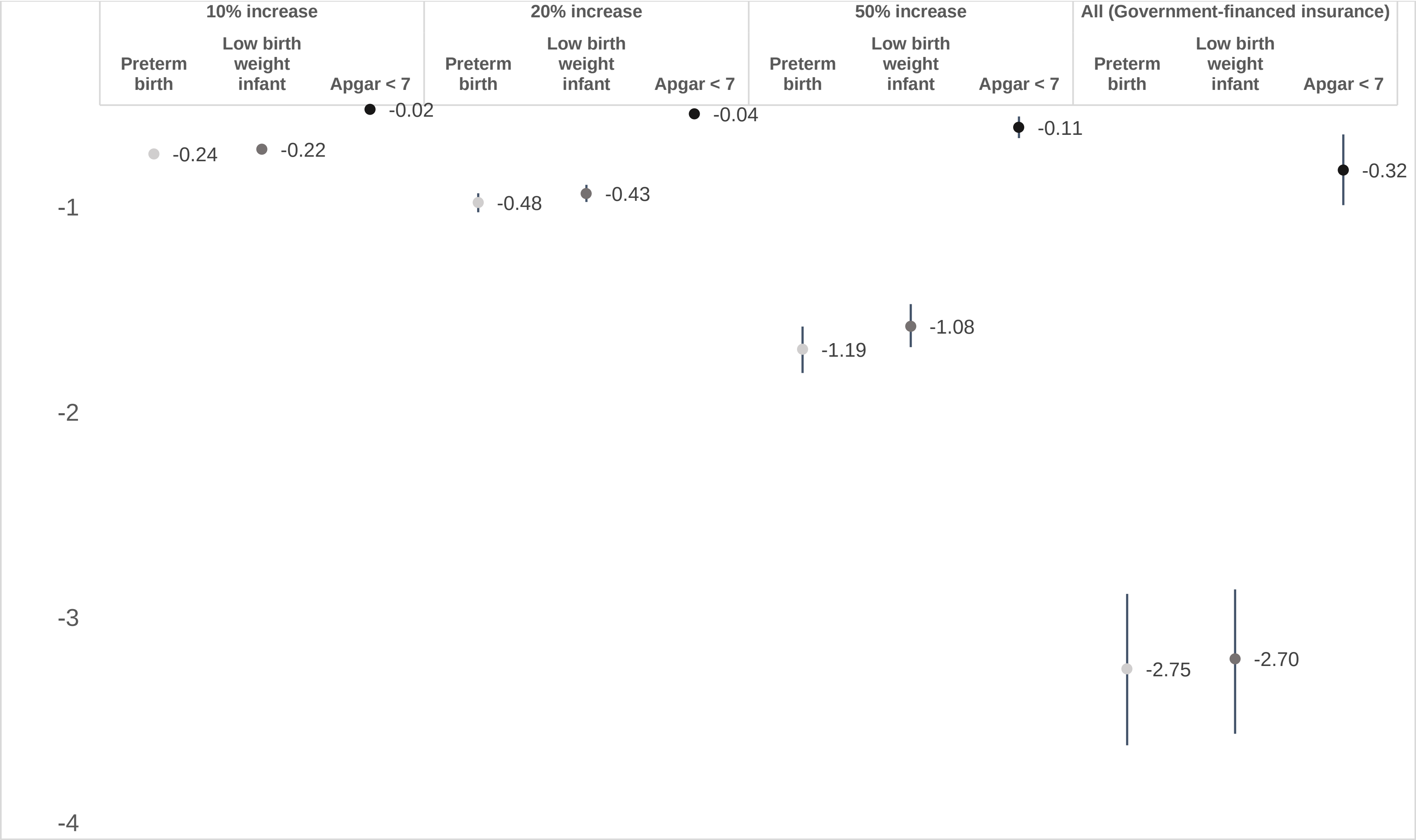

**Supplementary Figure 4. Change in prevalence of (a)cesarean section, labor induction, epidural and (b)postpartum hemorrhage, obstetric trauma, maternal infection stratified by parity with increase in use of CNMs as providers of prenatal care among low-risk pregnancies in Massachusetts, 2015-2019**

**(a)**

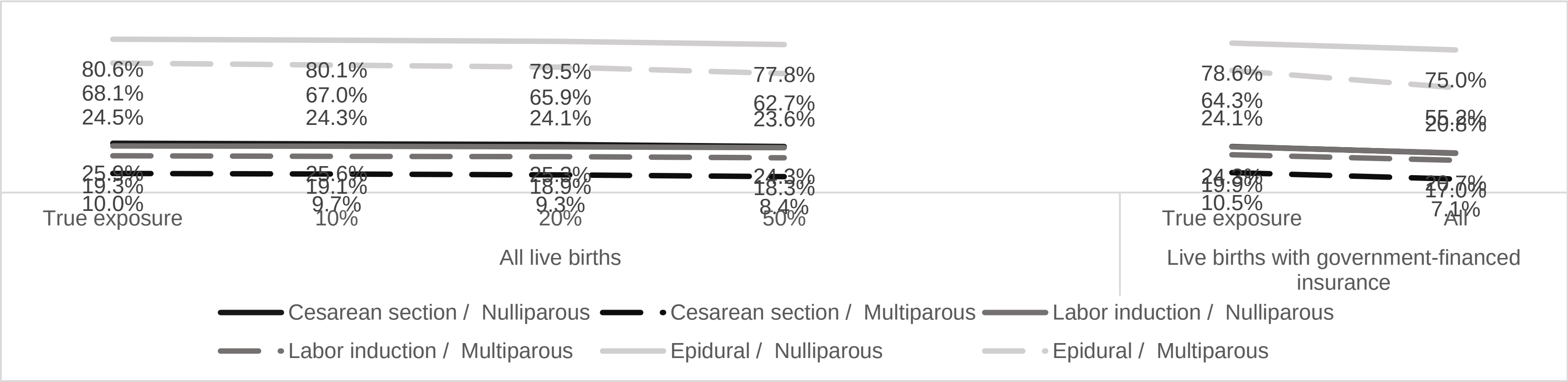


**(b)**

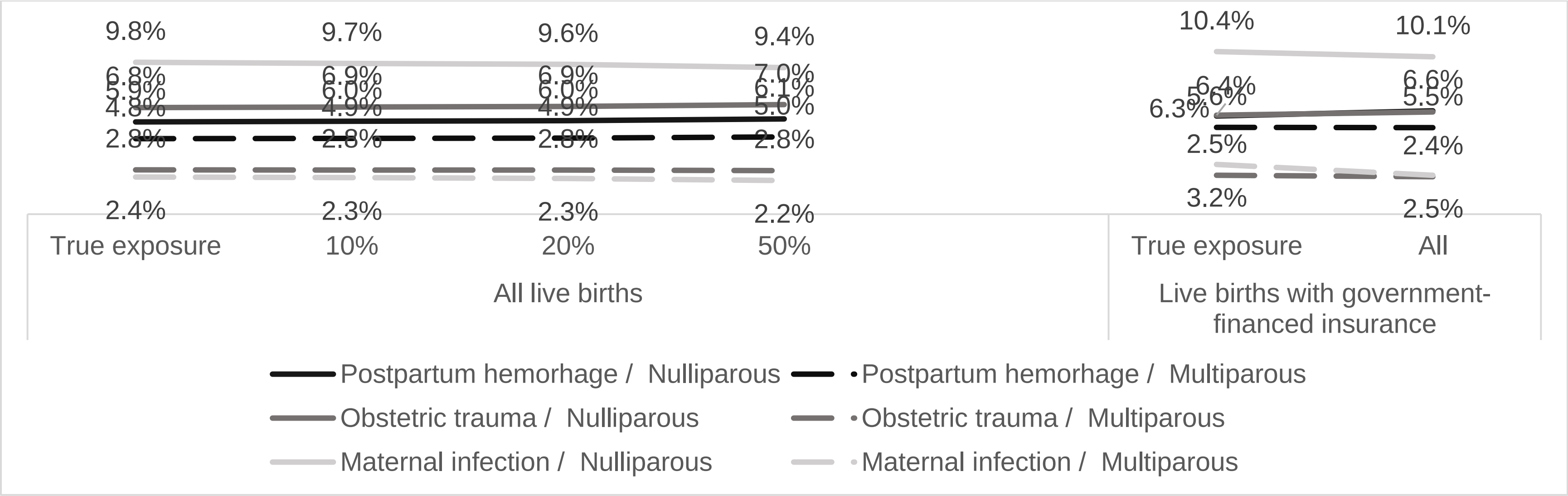

**Supplementary Figure 5. Prevalence difference of (a)cesarean section, labor induction, epidural and (b)postpartum hemorrhage, obstetric trauma, maternal infection stratified by parity with increase in use of CNMs as providers of prenatal care among low-risk pregnancies in Massachusetts, 2015-2019**

(a)

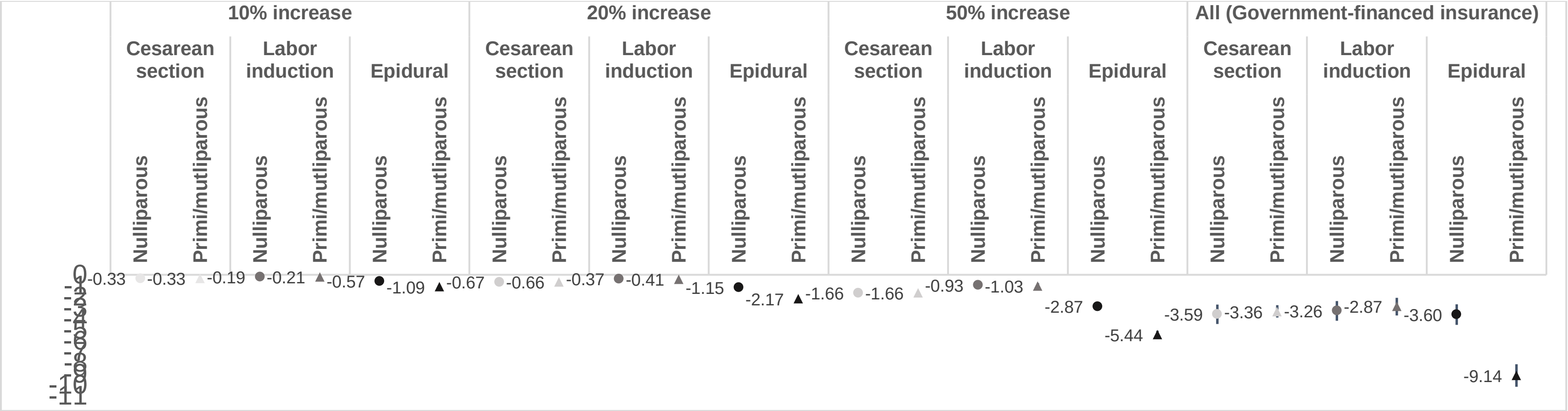


(b)

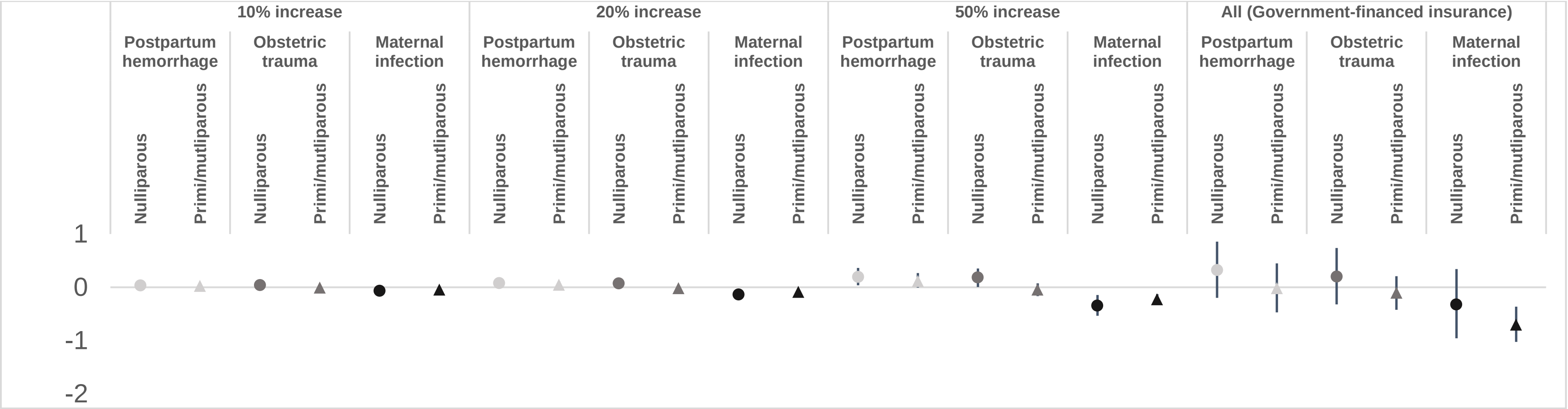

**Supplementary Figure 6. Prevalence (a) and prevalence difference (b) of preterm birth, low birth weight infants and infants with Apgar < 7 comparing increase in use of CNMs as prenatal care providers with natural course, stratified by parity, among low-risk pregnancies in Massachusetts, 2015-2019**

**(a)**

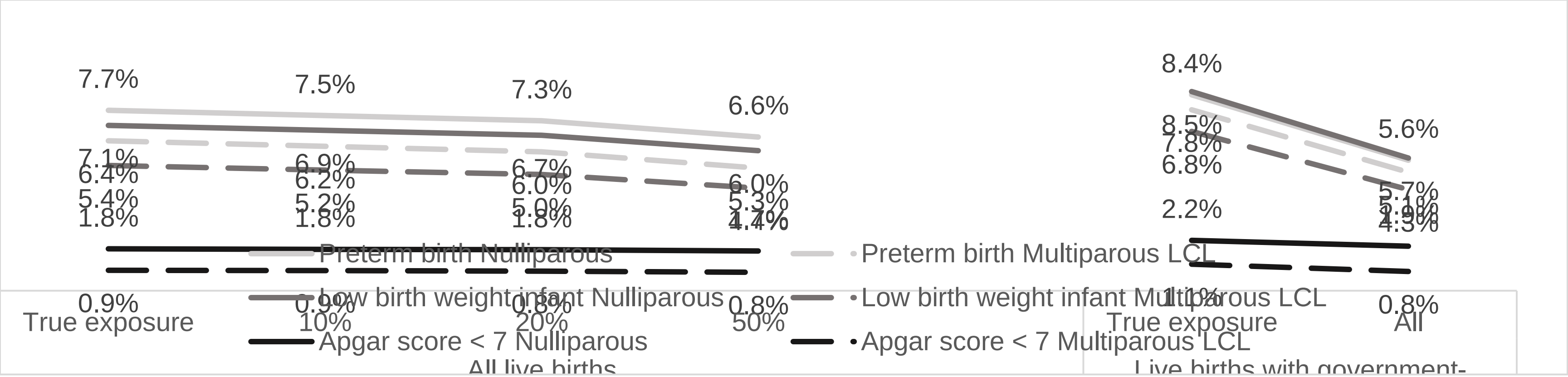


**(b)**

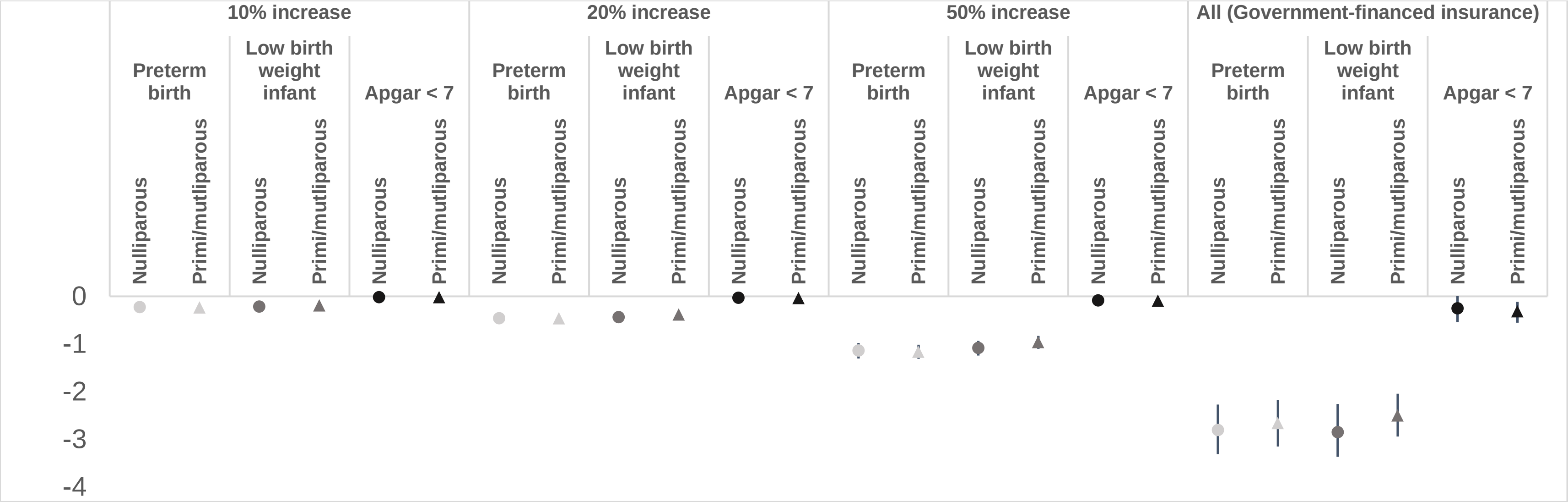

**Supplementary Figure 7. Prevalence (a) and prevalence difference (b) of postpartum hemorrhage comparing increasing use of CNMs as prenatal care providers with true exposure among low-risk pregnancies in Massachusetts, in 2015-2019, 2015-2016 and 2017-2019.**

**(a)**

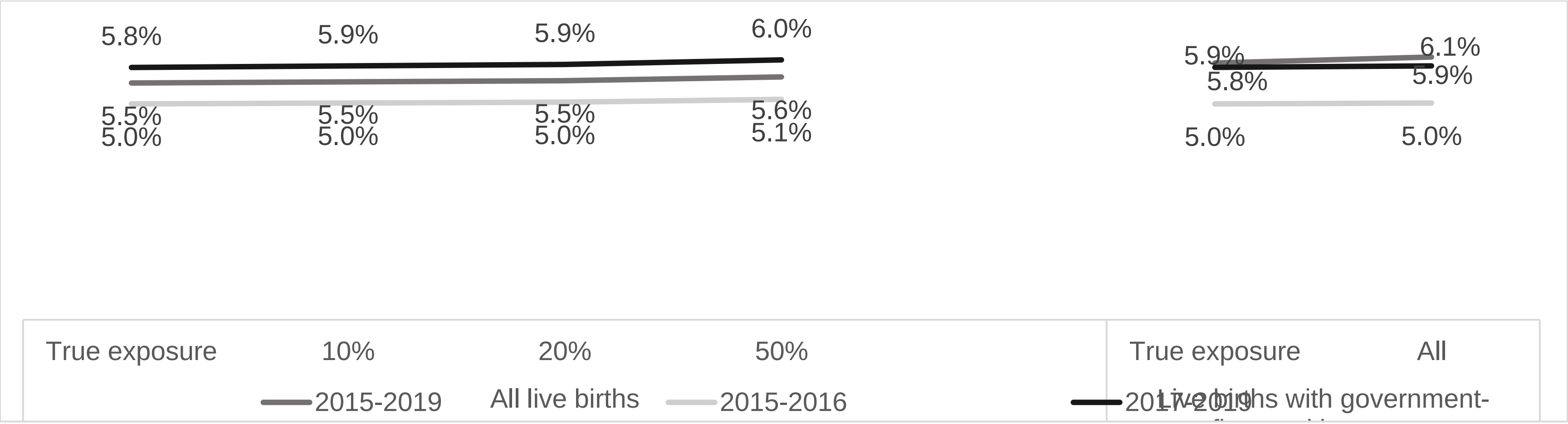


**(b)**

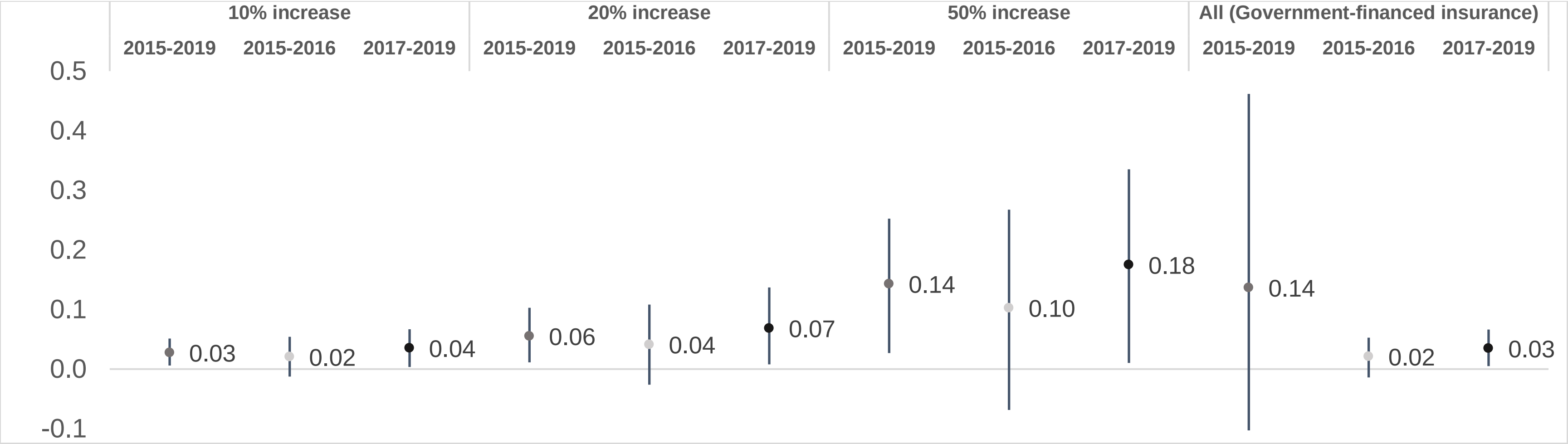

**Supplementary Figure 8. Prevalence (a) and prevalence difference (b) in postpartum hemorrhage and severe postpartum hemorrhage comparing increasing use of CNMs as prenatal care providers with true exposure among low-risk pregnancies in Massachusetts, 2015-2019**

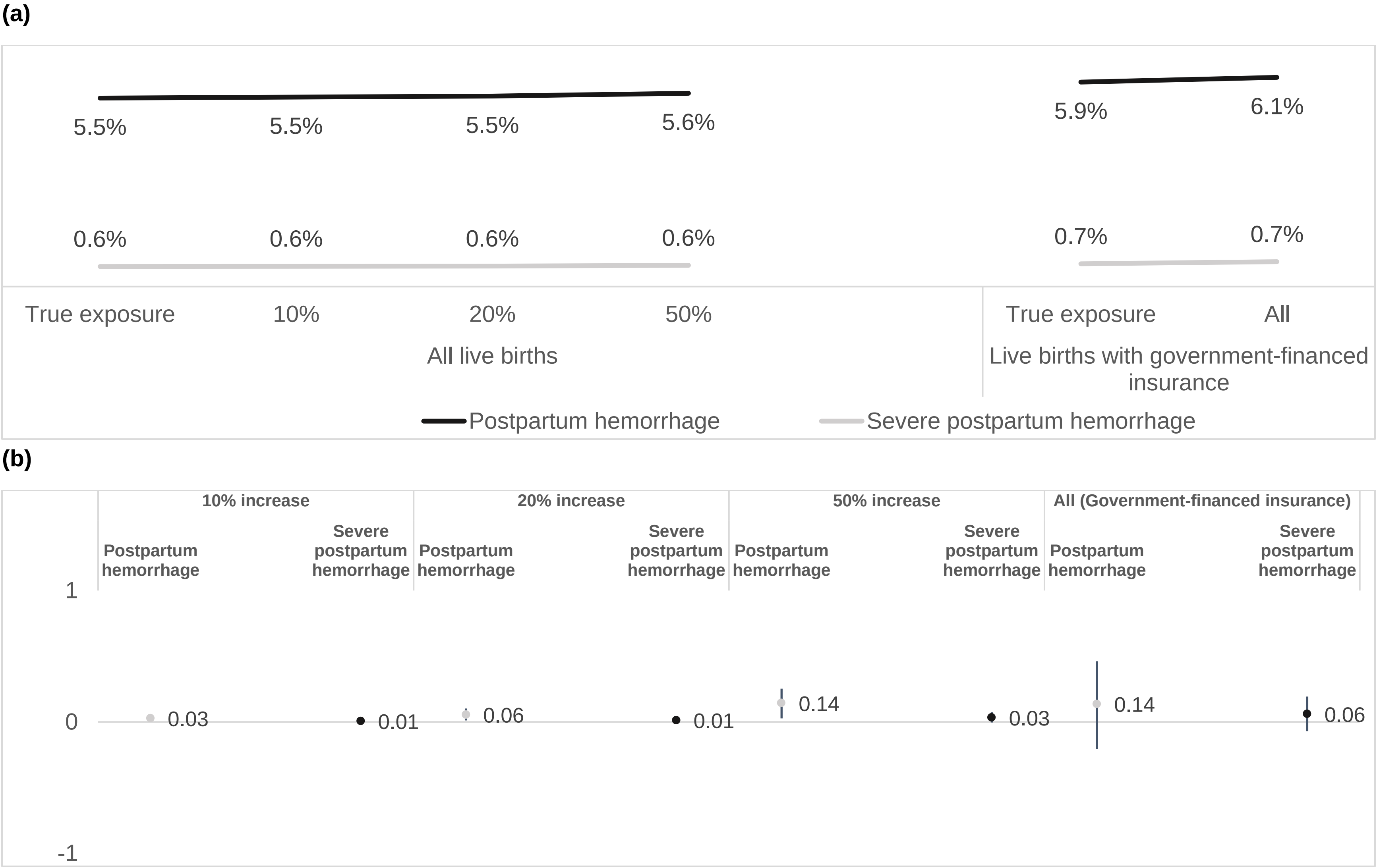
